# Adverse events following COVISHIELD (ChAdOx1nCoV-19) vaccination among health care workers in Sri Lanka; a multi-centre cross sectional survey

**DOI:** 10.1101/2022.01.26.22269788

**Authors:** Suranga Ravinda Manilgama, Nirosha Madhuwanthi Hettiarachchi, I Kumudini Jayasinghe, Shamila T De Silva, Thilak Jayalath, Thushari Wanigaratne, Bandusiri P Rathnayake, Navaneethakrishnan Suganthan, P Sudarshan, Manoji Pathirage, Nalayini Rajaratnam, Ganaka Senaratne, Vishaka Rajapaksha, Asanga Wickramasinghe, S P A L Ranaweera, Arjuna H M Thilakarathna, M Thilini D Kulaweera

**Affiliations:** Colombo North Teaching Hospital, Ragama; Teaching Hospital Peradeniya; National Hospital Kandy; Faculty of Medicine, University of Kelaniya; Faculty of Medicine, University of Peradeniya; District General Hospital, Matale; Teaching Hospital Rathnapura; Faculty of Medicine, University of Jaffna; District General Hospital, Polonnaruwa; Teaching Hospital Jaffna; Teaching Hospital Karapitiya; High School Advanced Mathematics, Gampaha

**Keywords:** COVISHIELD, ChAdOx1nCoV-19, Oxford/AstraZeneca, COVID-19 vaccination, adverse events, health care workers

## Abstract

**Introduction:** A community vaccination programme is the best approach to combat the COVID-19 pandemic. Post-vaccine surveillance is important to identify adverse events (AE) following COVID -19 vaccination in the population.

**Methods:** A multicentre cross-sectional survey was conducted in six provinces to estimate the prevalence of AE following the first dose of COVISHIELD (ChAdOx1nCoV-19) among all categories of health care workers (HCWs). A self-administered questionnaire was used to gather demographic data and AE.

**Results:** Of 5140 participants 67.8% were females. The mean (SD) age was 40.69 (±9.85) years. At least one comorbidity was reported in 15.4%. At least one AE was reported in 86.6% and 49.3% had local AE. Fever (67.2%), headaches (57.3%), body aches (54.4%), chills (51.2%), fatigue (37.5%) and arthralgia (36%) were the most reported systemic AE. The majority of AE lasted less than 24 hours. Pain and redness at the site were the most reported local AE. Mean duration of onset of fever and pain at injection site from the time of the vaccination was 6.65 and 9.67 hours respectively.

When participants were divided into two groups by mean age (≤40 and >40 years) and parameters were compared, most systemic (fever, nausea, fatigue, itching) and all local AE were significantly more prevalent in the ≤40 age group.

Two percent had reactions within the first 20 minutes. Anaphylaxis developed in 12 participants. Past history of anaphylaxis, drug or food allergy were reported in 0.6%, 2.8% and 6.7% respectively. However, previous history of allergy was not significantly related to immediate reactions or anaphylaxis following vaccination. Despite having minor AE, 71.1% attended routine work while 0.2% required hospitalisation.

**Conclusions:** While 86.6% reported minor AE, only a few serious AE were reported. Overall, the first dose of the vaccine was well-tolerated by HCWs.

## Introduction

The global pandemic of COVID-19 infection resulted in a range of clinical outcomes, varying from asymptomatic infection to severe acute respiratory distress and death (1, 2).

An effective vaccine is the best long-term answer to a pandemic. The development of vaccines against COVID-19 was fast-tracked due the availability of better technologies. A vaccine for a pandemic should protect the vulnerable population against severe disease and reduce deaths and transmission of the virus.

Sri Lanka received COVISHIELD (ChAdOx1nCoV-19 Coronavirus vaccine-recombinant) vaccine which is a product of the Serum Institute of India. It is the same vaccine as the Oxford/AstraZeneca vaccine. Priority in vaccination was given to healthcare workers and other frontline support staff by the COVID-19 task force.

Like most vaccines, there are adverse events ascribed to the vaccine by the manufacturer. Common AE following vaccination were described as local and systemic effects. Local AE included pain, warmth, redness, itching, and swelling at the injection site, and systemic AE included fatigue, chills, feeling feverish, fever, headache, nausea, joint pain, muscle pain, and body aches (3).

The vaccine development cycle includes preclinical, clinical (phase I, II and III) and manufacturing (phase IV) stages. During the clinical stages, safety, immunogenicity and efficacy is assessed in a stepwise manner. Comparative clinical studies and post-marketing surveillance are done during Phase IV (4). These steps are mandatory for a safe and swift immunisation programme. Similarly, a reporting mechanism needs to be in place for AE, which is both transparent and up to date.

Sri Lanka has a robust vaccination programme and a post-vaccine surveillance system for children. However, vaccination strategy against the pandemic was quite different and an adult vaccination programme of this magnitude has never been implemented previously. Since healthcare workers received the vaccine as a priority group, it was required to conduct a post-vaccine surveillance simultaneously among them. Therefore, this study was designed to identify common and important AE which can be monitored by any national AE reporting system in future.

The primary objective of the study was to assess the prevalence of major and minor AE among healthcare and ancillary workers at selected government hospitals in Sri Lanka. We intended to identify and describe the character, severity, and natural history of AE in the study population. The secondary objective was to assess socio-demographic factors and comorbidities associated with AE.

## Methods

This cross-sectional survey was carried out from March to May 2021 in eight selected government hospitals in the country, which were Colombo North Teaching Hospital, Teaching Hospital Peradeniya, National Hospital Kandy, District General Hospital Matale, Provincial General Hospital Polonnaruwa, Teaching Hospital Ratnapura, Teaching Hospital Karapitiya, Teaching Hospital Jaffna.

All categories of health care workers (HCWs) and ancillary staff members who received the first dose of the COVISHIELD vaccine and consented to participate in the study were included. Any HCW who received any other type of vaccine was excluded. HCWs who received the vaccine at the participating government hospitals were approached within two weeks of vaccination to gather information on vaccine related AE. After obtaining informed written consent participants were asked to fill the study questionnaire in their preferred language of Sinhala, English, or Tamil. The study questionnaire consisted of the following components: demographic data, special circumstances such as pregnancy and breastfeeding, history of previous COVID infection, previous allergic history, presence of comorbidities, immediate adverse reactions (within first 20 minutes), details of systemic and local AE, and outcome following vaccination.

The severity of AE, and the onset and duration of each adverse reaction were documented. Allergic reactions such as anaphylaxis, angioedema, itching, urticaria, wheezing or skin rashes were included. Outcome of AE was recorded to identify any work abstinence, hospitalisation for management of AE, and disability.

Statistical Package for Social Sciences (SPSS) software was used for the analysis of data. Descriptive statistics were used to describe population parameters. Prevalence of various AE in the total study population and subgroups were calculated. Factors associated with binary outcomes (e.g., presence or absence of significant comorbidities) were assessed using multiple logistic regression, adjusting for confounding variables.

## Results

Overall, 5140 health care workers completed the questionnaire. The mean (SD) age of participants was 40.69 (±9.85) years; 67.8% were females. The composition was 17.6% medical doctors, 36.4% nurses, and 22.9% health care assistants. Paramedics and other supportive staff comprised the rest (Table 1). Only 1% (n=52) had tested positive for SARS-CoV2 PCR previously. Breastfeeding mothers represented 2.1% (n=110) of the study population while 0.14% (n=7) were pregnant.

**Table 1.** Composition of the study population

| **Gender** | **Percentage (%)** | **Number (n=)** |
| --- | --- | --- |
| Male | 32.2 | 1646 |
| Female | 67.8 | 3469 |
| **Ethnicity** |  |  |
| Sinhalese | 76.5 | 3873 |
| Others | 23.5 | 1187 |
| **Health care worker category** |  |  |
| Medical | 17.6 | 898 |
| Nursing | 36.4 | 1855 |
| Health care assistants | 22.9 | 1169 |
| Professions supplementary to medicine | 4.2 | 216 |
| Paramedical staff | 2.7 | 135 |
| Hospital office staff | 3.5 | 179 |
| Security officers | 4.5 | 230 |
| Hospital cleaning staff | 4.8 | 246 |
| Ambulance staff | 0.5 | 24 |
| Others | 2.7 | 143 |
| Pregnant | 0.14 | 7 |
| Breast feeding | 2.1 | 110 |
| Past infection (positive PCR for SAR-CoV2) | 1 | 52 |

Distribution of comorbidities among participants and previous history of allergy is shown in Table 2. At least one comorbidity was reported in 18.8%. The presence of comorbidities increased with advancing age. The most prevalent comorbidities were bronchial asthma, diabetes, and hypertension at 5.7%, 5.4% and 4.5% respectively. The associations of the presence of comorbidities and the occurrence of both local and systemic adverse events were analysed separately and collectively, but they were not significant. A history of allergy was present in 13.3% of the study population while only 0.6% had a history of anaphylaxis. 6.7% had a food allergy, 2.8% had a drug allergy and 1.1% had a previous vaccine-related adverse event.

**Table 2.**
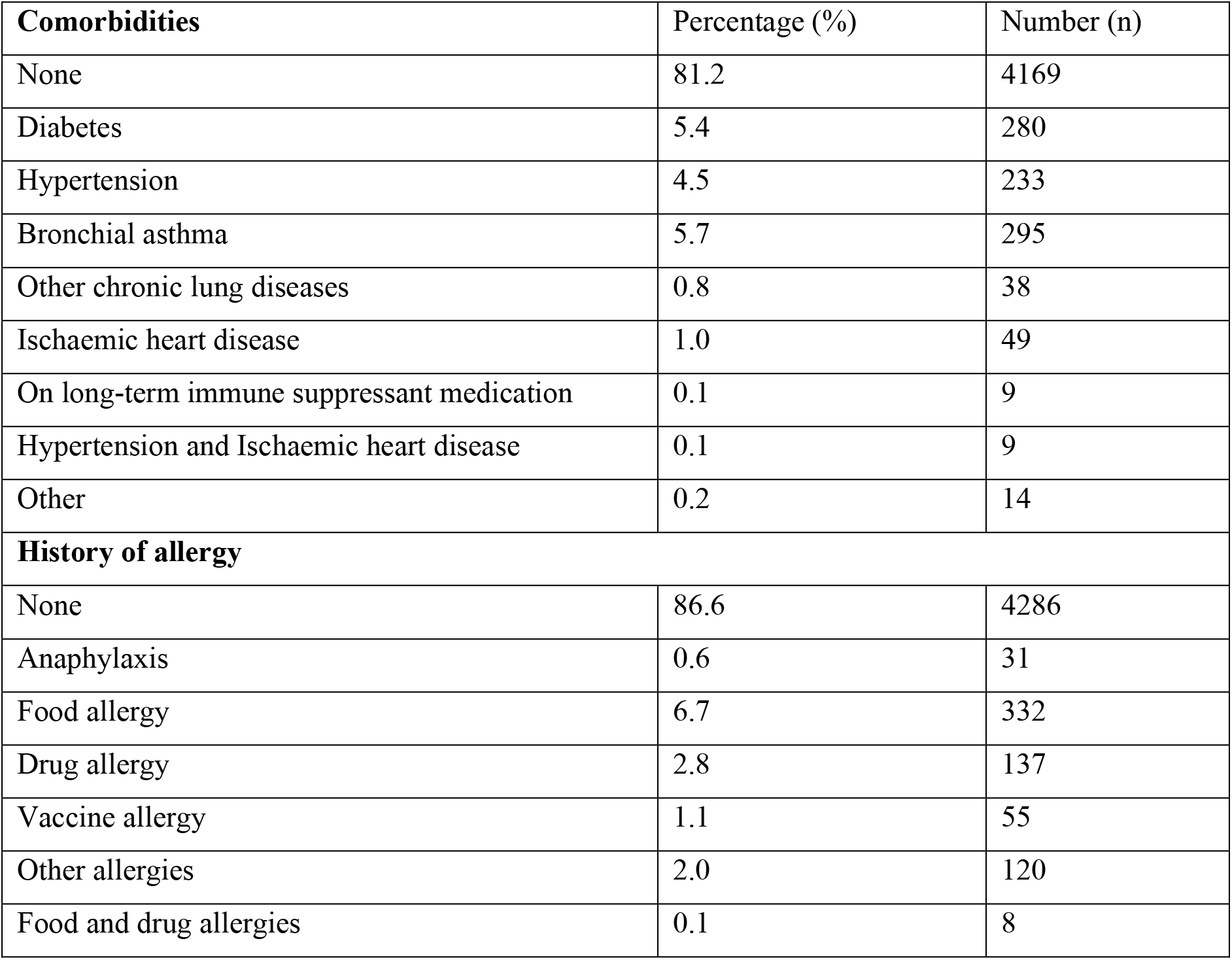
Distribution of comorbidities and history of allergy in the study population

The study population was divided into two groups using the mean age of the population; individuals aged ≤ 40 years (younger age group) and individuals aged >40 years (older age group). Variables were compared between the two groups. Categorical variables were compared using chi square test and continuous variables were compared using student-t test. AE following vaccination were categorised as immediate (within 20 minutes of vaccination, which is the duration of observation at the vaccination centre) and late (20 minutes after vaccination). Two percent of vaccine recipients developed AE within the first 20 minutes. Anaphylaxis developed in 12 patients (major immediate AE), and all were treated successfully. 0.8% developed urticaria. A history of anaphylaxis, drug, or food allergy did not have a significant relation to current vaccine induced immediate reactions or anaphylaxis. Interestingly, none of the individuals who developed anaphylaxis after the current vaccine had a history of anaphylaxis.

Overall, 86.6% of vaccine recipients had at least one AE and 49.3% of vaccine recipients had at least one local AE. Systemic and local AE that were reported are shown in Table 3. Fever was recorded as low-grade (temperature 99-100.4°F) and high-grade (temperature >100.4°F). Participants also reported feeling “feverish” when there was a feeling of having fever but no increase in body temperature. Those aged 40 years and younger were significantly more likely to experience feeling feverish, rigors, and vomiting (p<0.02).

**Table 3.**
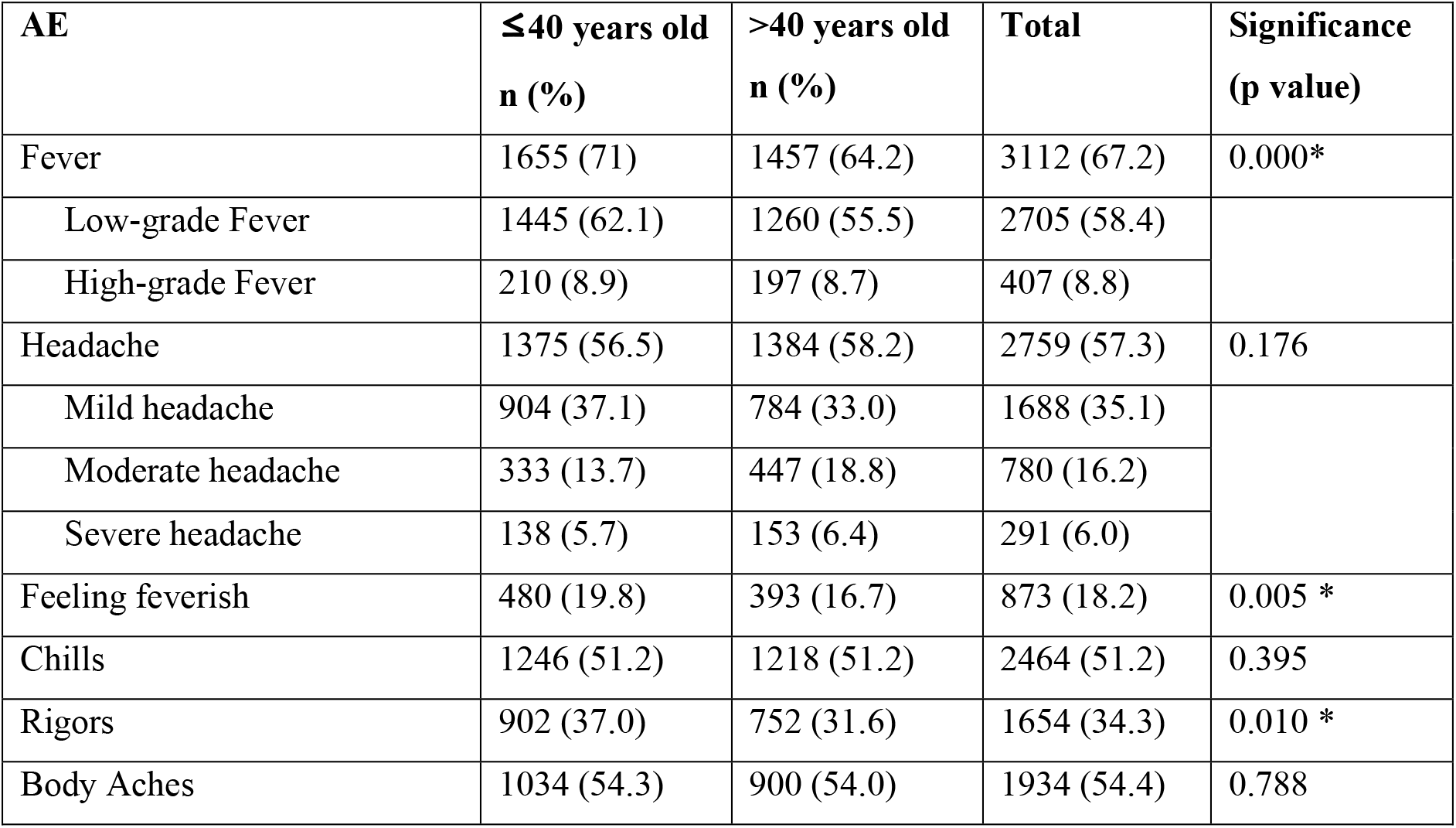
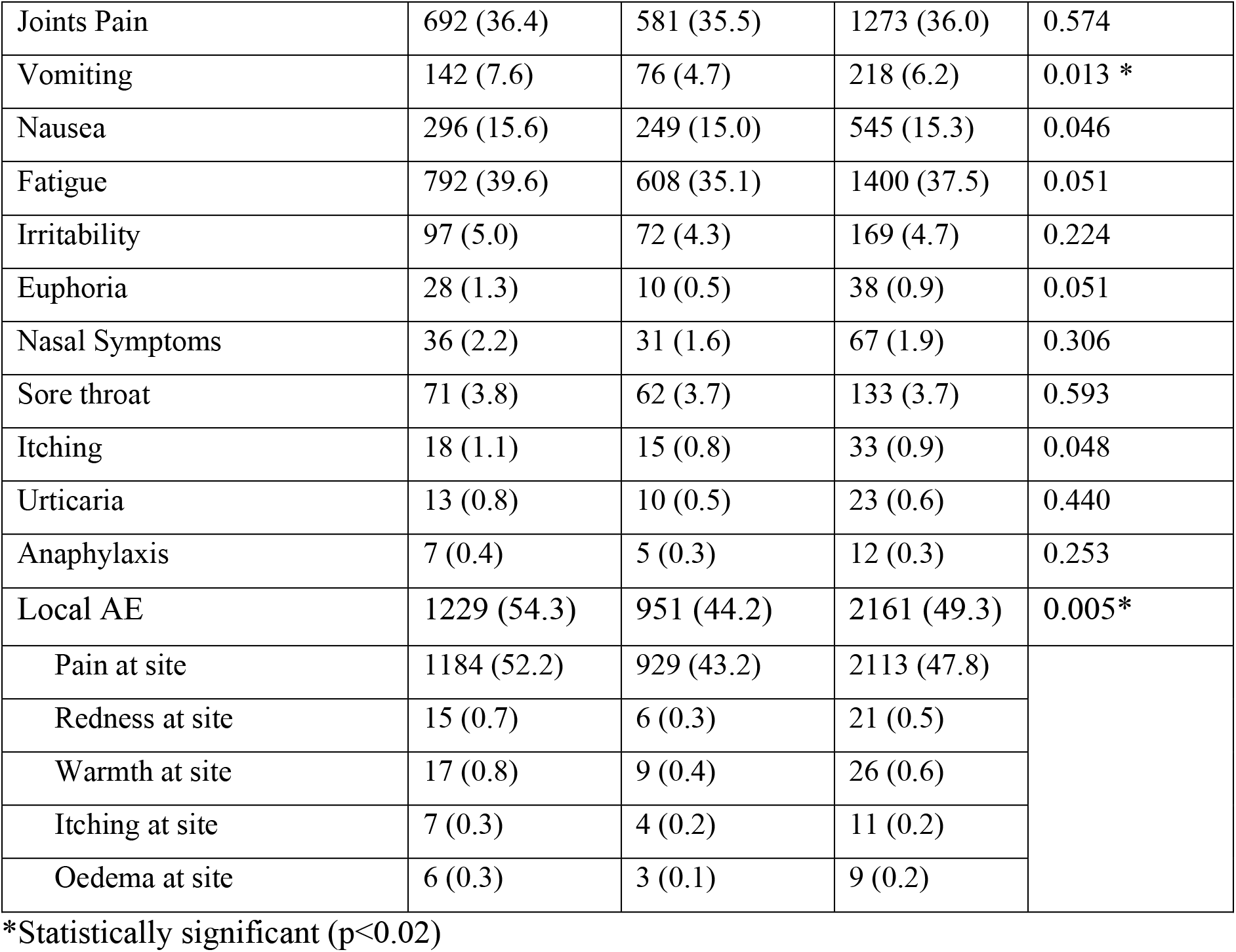
Occurrence of systemic and local AE among vaccine recipients

| AE | ≤40 years old<br>n (%) | >40 years old<br>n (%) | Total | Significance<br>(p value) |
| --- | --- | --- | --- | --- |
| Fever | 1655 (71) | 1457 (64.2) | 3112 (67.2) | 0.000* |
| Low-grade Fever | 1445 (62.1) | 1260 (55.5) | 2705 (58.4) |  |
| High-grade Fever | 210 (8.9) | 197 (8.7) | 407 (8.8) |  |
| Headache | 1375 (56.5) | 1384 (58.2) | 2759 (57.3) | 0.176 |
| Mild headache | 904 (37.1) | 784 (33.0) | 1688 (35.1) |  |
| Moderate headache | 333 (13.7) | 447 (18.8) | 780 (16.2) |  |
| Severe headache | 138 (5.7) | 153 (6.4) | 291 (6.0) |  |
| Feeling feverish | 480 (19.8) | 393 (16.7) | 873 (18.2) | 0.005 * |
| Chills | 1246 (51.2) | 1218 (51.2) | 2464 (51.2) | 0.395 |
| Rigors | 902 (37.0) | 752 (31.6) | 1654 (34.3) | 0.010 * |
| Body Aches | 1034 (54.3) | 900 (54.0) | 1934 (54.4) | 0.788 |
| Joints Pain | 692 (36.4) | 581 (35.5) | 1273 (36.0) | 0.574 |
| Vomiting | 142 (7.6) | 76 (4.7) | 218 (6.2) | 0.013 * |
| Nausea | 296 (15.6) | 249 (15.0) | 545 (15.3) | 0.046 |
| Fatigue | 792 (39.6) | 608 (35.1) | 1400 (37.5) | 0.051 |
| Irritability | 97 (5.0) | 72 (4.3) | 169 (4.7) | 0.224 |
| Euphoria | 28 (1.3) | 10 (0.5) | 38 (0.9) | 0.051 |
| Nasal Symptoms | 36 (2.2) | 31 (1.6) | 67 (1.9) | 0.306 |
| Sore throat | 71 (3.8) | 62 (3.7) | 133 (3.7) | 0.593 |
| Itching | 18 (1.1) | 15 (0.8) | 33 (0.9) | 0.048 |
| Urticaria | 13 (0.8) | 10 (0.5) | 23 (0.6) | 0.440 |
| Anaphylaxis | 7 (0.4) | 5 (0.3) | 12 (0.3) | 0.253 |
| Local AE | 1229 (54.3) | 951 (44.2) | 2161 (49.3) | 0.005* |
| Pain at site | 1184 (52.2) | 929 (43.2) | 2113 (47.8) |  |
| Redness at site | 15 (0.7) | 6 (0.3) | 21 (0.5) |  |
| Warmth at site | 17 (0.8) | 9 (0.4) | 26 (0.6) |  |
| Itching at site | 7 (0.3) | 4 (0.2) | 11 (0.2) |  |
| Oedema at site | 6 (0.3) | 3 (0.1) | 9 (0.2) |  |

\*Statistically significant (p<0.02)

Pain at the injection site (47.8%) was the most reported local AE with almost half the study population experiencing the symptom. All localised AE were common in the younger age group and there was a significant difference between the two age groups (p<0.02).

Table 4 shows the onset of AE following vaccination in hours. Fever, headache, body aches, arthralgia, itching, and fatigue developed significantly earlier in the younger age group.

**Table 4.**
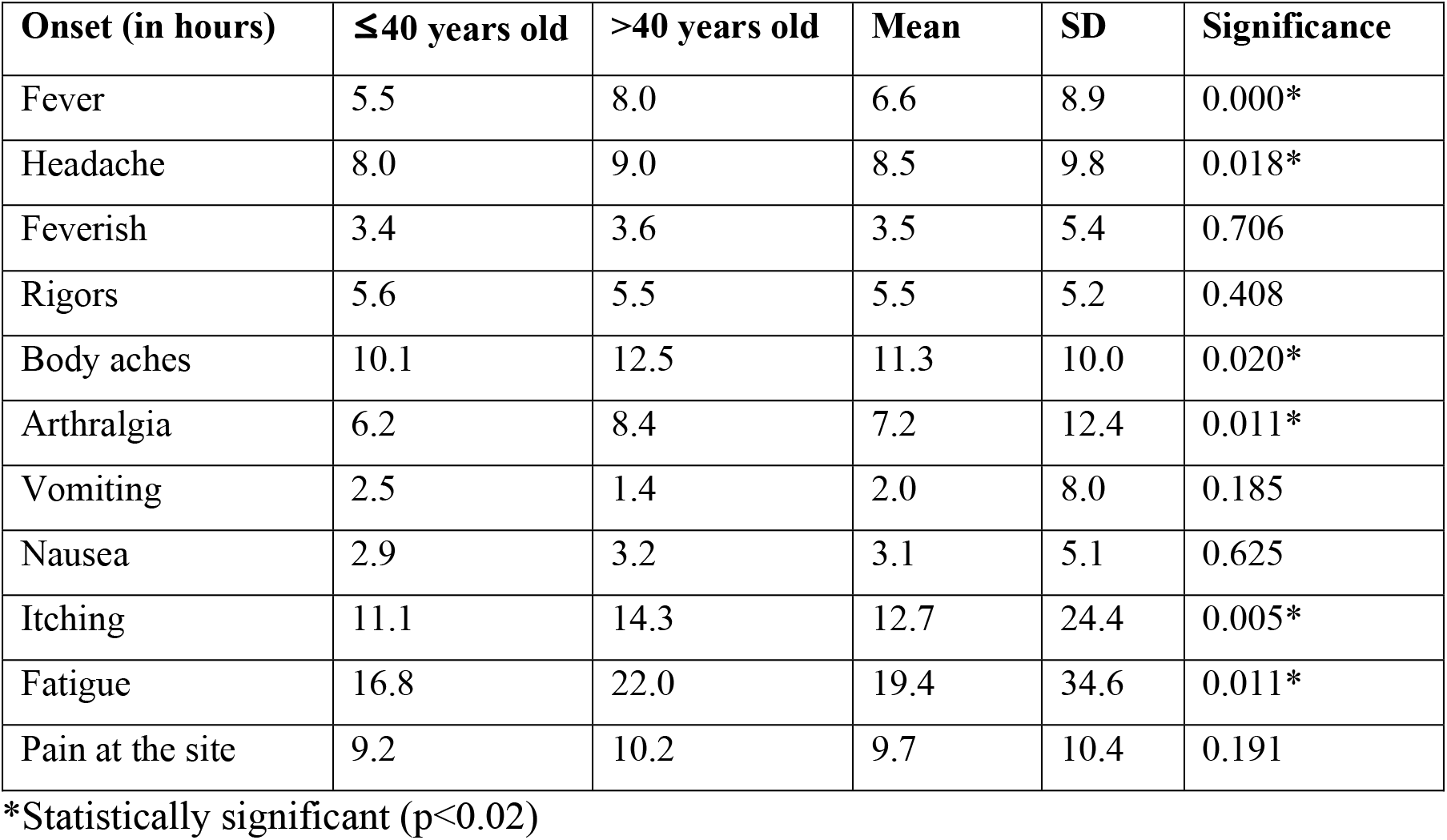
Onset of adverse events following vaccination (in hours)

The duration of AE was analysed (Table 5). A majority of AE lasted less than 24 hours. Fever, headache, body aches and arthralgia lasted longer in those over 40 years of age.

**Table 5.** Duration of adverse events following vaccination (in hours)

| Duration (in hours) | ≤40 years old | >40 years old | Mean | SD | Significance |
| --- | --- | --- | --- | --- | --- |
| Fever | 14.6 | 20.2 | 17.1 | 21.8 | 0.000* |
| Headache | 23.8 | 28.3 | 26.1 | 35.1 | 0.005* |
| Feverish | 10.5 | 11.4 | 10.9 | 19.2 | 0.449 |
| Rigors | 10.9 | 11.8 | 11.3 | 18.6 | 0.376 |
| body aches | 16.1 | 19.1 | 17.7 | 43.6 | 0.043 |
| Arthralgia | 13.2 | 17.4 | 15.4 | 52.8 | 0.045 |
| Vomiting | 3.7 | 4.2 | 4.0 | 14.2 | 0.655 |
| Nausea | 12.7 | 14.4 | 13.7 | 36.1 | 0.056 |
| Fatigue | 19.4 | 18.9 | 19.3 | 60.7 | 0.415 |

\*Statistically significant (p<0.02)

Outcome of AE is shown in Table 6. The majority (71.1%) attended routine work despite having minor AE and only a small number required hospitalisation. The vaccination programme was conducted close to the weekends in most hospitals. Some of the HCWs scheduled their vaccination prior to their off days anticipating AE. Therefore, 24.0% of vaccine recipients stayed home and rested for 1-2 days following the vaccination. Outcome of the AE in the older age group was favourable and showed a significant difference across age groups (p=0.000).

**Table 6.** Outcome of AE among HCWs

| Outcome | ≤40 years old<br>n (%) | >40 years old<br>n (%) | Total<br>n (%) |
| --- | --- | --- | --- |
| Attended routine work | 1617 (68.3) | 1715 (72.8) | 3332 (71.1) |
| Stayed home and rested | 572 (24.5) | 553 (23.5) | 1125 (24.0) |
| Admitted and observed | 7 (0.25) | 4 (0.15) | 11 (0.2) |
| Admitted and treated | 5 (0.2) | 3 (0.1) | 8 (0.1) |
| Off duty (sick leave) | 135 (5.8) | 77 (3.3) | 212 (4.5) |

## Discussion

In this multicentre cross-sectional survey of adverse events following the first dose of the COVISHIELD (ChAdOx1nCoV-19) vaccination among health care workers in Sri Lanka, at least one AE was reported in 86.6% and 49.3% had local AE. Fever (67.2%), headaches (57.3%), body aches (54.4%) and chills (51.2) were the most reported systemic AE. The majority of AE lasted less than 24 hours. Pain and redness at the site were the most reported local AE. Mean duration of onset of fever and pain at injection site was 6.65 and 9.67 hours respectively. Most systemic (fever, nausea, fatigue, itching) and all local AE were significantly more prevalent in the ≤40 age group. While 2% percent had reactions within the first 20 minutes, anaphylaxis developed in 12 participants; a history of allergy was not significantly related to immediate reactions or anaphylaxis following vaccination. Despite having minor AE, 71.1% attended routine work while 0.2% required hospitalisation. Though 86.6% reported minor AE there were only a few serious AE. Overall, the first dose of the vaccine was well-tolerated by HCWs.

Our study was conducted among HCWs in Sri Lanka, at a time when there were no large-scale post-vaccination surveillance studies. The mean age of the study population was 40.69 (±9.85) years, with an age range of 18-63 years. The study population was divided into two groups, ≤40 years old and >40 years old, considering the mean age. The profiles of adverse events were compared between the two groups. Two-thirds of the sample was female which reflected the increased proportion of female HCWs in government hospitals in the country.

With the introduction of the vaccination programme to Sri Lanka, priority was given to frontline healthcare workers and other stakeholders. This provided the ideal setup to study adverse events among frontline workers before it reached the public.

A national vaccination programme needs a safe vaccine and post-vaccine surveillance as an integral part of the programme. When a vaccine is introduced for a newly emerged disease there are standard steps to follow. Once it is recommended by regulatory authorities to be used in human beings, the post vaccination surveillance is an integral part of the whole cycle. Not only the target antigen, but also the other constituents like vector, stabiliser, adjuvants, and preservatives can give rise to various effects in the vaccine recipient. Those adverse effects can vary according to diverse factors in the population. Some of the adverse effects may be related to specific characteristics of the individual. Epigenetics of the target population, prior exposure and sensitization to similar vaccines and similar viral particles are some of the important factors that can affect outcome. Therefore, a vaccine may behave differently between two different populations, not only in its effectiveness but also in its adverse effects profile.

Our study revealed that the vaccine was well tolerated with less AE in the older age group (>40 years) compared to the <40-year-olds. AE after vaccination were mild to moderate in nature, which is similar to findings from studies elsewhere (5). At the initial phase of vaccine development, a single-blind, randomised controlled trial was conducted at five trial sites in the UK. Participants were assigned to receive either ChAdOx1 nCoV-19 vaccine (n=543) or meningococcal group A, C, W-135, and Y conjugate vaccine (n=534). Some received paracetamol prophylaxis. Median age of participants was 35 years (6) and fatigue and headache were the most reported systemic reactions. Fatigue was reported in 340 (70%) participants without paracetamol and in 40 (71%) with paracetamol. Headaches were reported in 331 (68%) participants without paracetamol and in 34 (61%) with paracetamol. Other systemic adverse reactions reported in subgroups without paracetamol vs with paracetamol were muscle ache in 294 (60%) vs 27 (48%); chills in 272 (56%) vs 15 (27%); feeling feverish in 250 (51%) vs 20 (36%); temperature of at least 38°C in 87 (18%) vs 9 (16%); temperature of at least 39°C in 8 (2%) vs none. Pain at the site was reported in 67% and 50% of individuals without and with paracetamol prophylaxis respectively.

In our study fever (67.2%) and headache (57.3%) were the most reported systemic AE. Fatigue was reported in 37.5%, body aches in 54.4%, arthralgias in 36%, chills in 51.2% and feeling feverish in 18.2% of participants. Low-grade fever in 58.4% and high-grade fever in 8.8% of individuals were reported. Pain at the site was reported in 47.8% of individuals.

A study done in Bangladesh showed similar results for local AE, with pain at the site in 48.9%. But the occurrences of systemic AE were much lower, with fever and headache in 24.3% and 13.7% respectively (7). A study from South Africa reported fatigue (87.6%), myalgia (80.8%), headache (72.0%), and fever of ≥ 38.0°C (38.7%) as the most common adverse events among 5930 HCWs who received the first dose of AstraZeneca vaccine, but most symptoms resolved within 2 days (8).

In our study anaphylaxis developed in 12 persons (major immediate AE), and all were managed successfully. It is noteworthy that a history of anaphylaxis, drug, or food allergy did not have a significant relation to current vaccine-induced allergy or anaphylaxis. In our study population, although comorbidities showed a significant difference across age groups, there was no significant association between having a comorbidity and developing systemic or local AEs.

Several systemic (fever, feverishness, rigors, vomiting) and all local AE were significantly more likely in the younger age group compared to the older age group. Similarly, a study done in HCWs in Togo following the first dose of COVISHIELD vaccine found that prevalence of AE was significantly more common in the younger age group (9). A Study done in Bangladesh also revealed a similar trend where presence and number of AEs were significantly greater in younger adults (7).

In our study, the onset of AE was earlier in the younger age group, but AEs lasted longer in the older age group. Fever and headache developed earlier in the younger age group, while both lasted longer in the older age group. Ramasamy et al. has also reported that ChAdOx1 nCoV-19 appears to be better tolerated in older people compared to younger adults (5).

Since the introduction of vaccination, there have been concerns about unexpected adverse events. Four randomised controlled trials conducted to assess the safety and efficacy of ChAdOx1 nCoV-19 vaccine in three countries reported the possible association of vaccination with serious adverse events such as transverse myelitis (10). Concerns have been raised about possible COVISHIELD vaccine induced thromboembolic events. Analysis of thrombotic adverse reactions to COVISHIELD vaccine reported to EudraVigilance database from 17^th^ February to 12^th^ March 2021 identified 28 thrombotic events linked to vaccination out of 54,571 adverse events, 3 of which were fatal outcomes of pulmonary embolism (PE) (11). However, none of our study participants experienced any neurological or thrombotic complications.

### Strengths and limitations

Recall bias must be considered when documenting onset time and duration of AE. Validated scales are ideal when assessing severity and outcome of AE in large populations. Our study was helpful in identifying the profile of AE following vaccination in a local population in the absence of post-vaccination surveillance in the country.

### Conclusions and recommendations

Those younger than 40 years reported a higher number of most of the systemic and all local AE. Almost four-fifth of the study population reported minor AE. Serious AE were very few; there were no thrombotic events or neurological complications. The first dose of the vaccine was well-tolerated by the majority of HCWs. Community based vaccine surveillance is mandatory to identify new onset AE following COVID-19 vaccination. The establishment of such a system as part of the routine COVID vaccination programme is strongly recommended.

## Data Availability

All data produced in the present work are contained in the manuscript

## Author contributions

Authors SRM, NMH, IKJ and STD conceptualised the study. SRM, NMH, IKJ, STD, TJ, TW, RPB, NS, PS, MP, NR, GS, VR, AW, SPALR, and HMAT were involved in data collection. MTDK was the bio statistician. All authors were involved in drafting the paper and revising the manuscript.

